# A fearful adult attachment style is associated with double the prevalence of chronic pain compared to secure attachment: A national survey of a South African population

**DOI:** 10.1101/2023.07.27.23293239

**Authors:** Gabriella Elisabeth Stamp, Stella Iacovides, Antonia Louise Wadley

## Abstract

Our response to threats, including pain, are believed to be learnt during our early interpersonal relationships, and can be measured through attachment style. Preliminary epidemiological evidence suggests that insecure attachment styles are more prevalent in those with chronic pain. Our aim was to determine the association between adult attachment style and chronic pain prevalence and burden in a South African population. A nationwide online survey determined adult attachment style (using The Experience in Close Relationships - Relationship Structures (ECR-RS) Questionnaire), prevalence of chronic pain and typically-associated psychological factors. In those with chronic pain, the association with attachment style and pain burden (pain sites, severity and interference, using the Brief Pain Inventory) was further determined. Results of the 2371 individuals were analyzed using multivariate generalized linear models. In our young (median age 23 years; IQR 20-28), well-educated and primarily female (74%) cohort with predominantly a middle-to-high socioeconomic status, we found a high prevalence of chronic pain (27%). All three insecure attachment styles were associated with increased chronic pain prevalence when compared to the secure attachment style (Dismissing: 31%, Odds ratio [95%CI] = 1.38 [1.02-1.85], p=0.037; Preoccupied: 42%, Odds ratio [95%CI] = 2.26 [1.62-3.13], p<0.001; Fearful: 49%, Odds ratio [95%CI] = 2.95 [2.03-4.29], p<0.001). Adult attachment style was not directly associated with the burden of chronic pain, because pain catastrophizing mediated this relationship. Adult attachment style was, however, directly associated with chronic pain prevalence, with more than double the chronic pain prevalence in the fearfully, compared to securely, attached individuals.

## Introduction

The impact of interpersonal relationships on whether a stimulus is perceived as threatening or safe is a key component of the social modulation of pain ^33^. The perceived intensity of an experimentally-induced painful stimulus can increase if a social threat is added, despite the experimental stimulus staying the same ^31, 46^. Conversely, the presence of a supportive other, whether imagined through the presence of a picture, or physically present and holding a hand, can reduce pain intensity and pain-related neural activity on functional magnetic resonance imaging (fMRI), again, even as the stimulus remains at the same intensity ^20, 52^.

Perceived safety or threat in various situations is likely predetermined by individuals’ prior learning and experiences. Whilst learning about pain and/or threat can take place throughout life, learning about the inherent safety or threat of the world begins in infancy through interactions with one’s primary caregiver ^8, 42^. This particular learning about inherent safety or threat gives rise to attachment behaviours, which can be classified into attachment styles. Essentially, these attachment styles represent an individual’s belief in themselves to cope, and their belief in others to support them, during times of stress (illustrated in Fig. 1) ^42^.

**Fig. 1:**
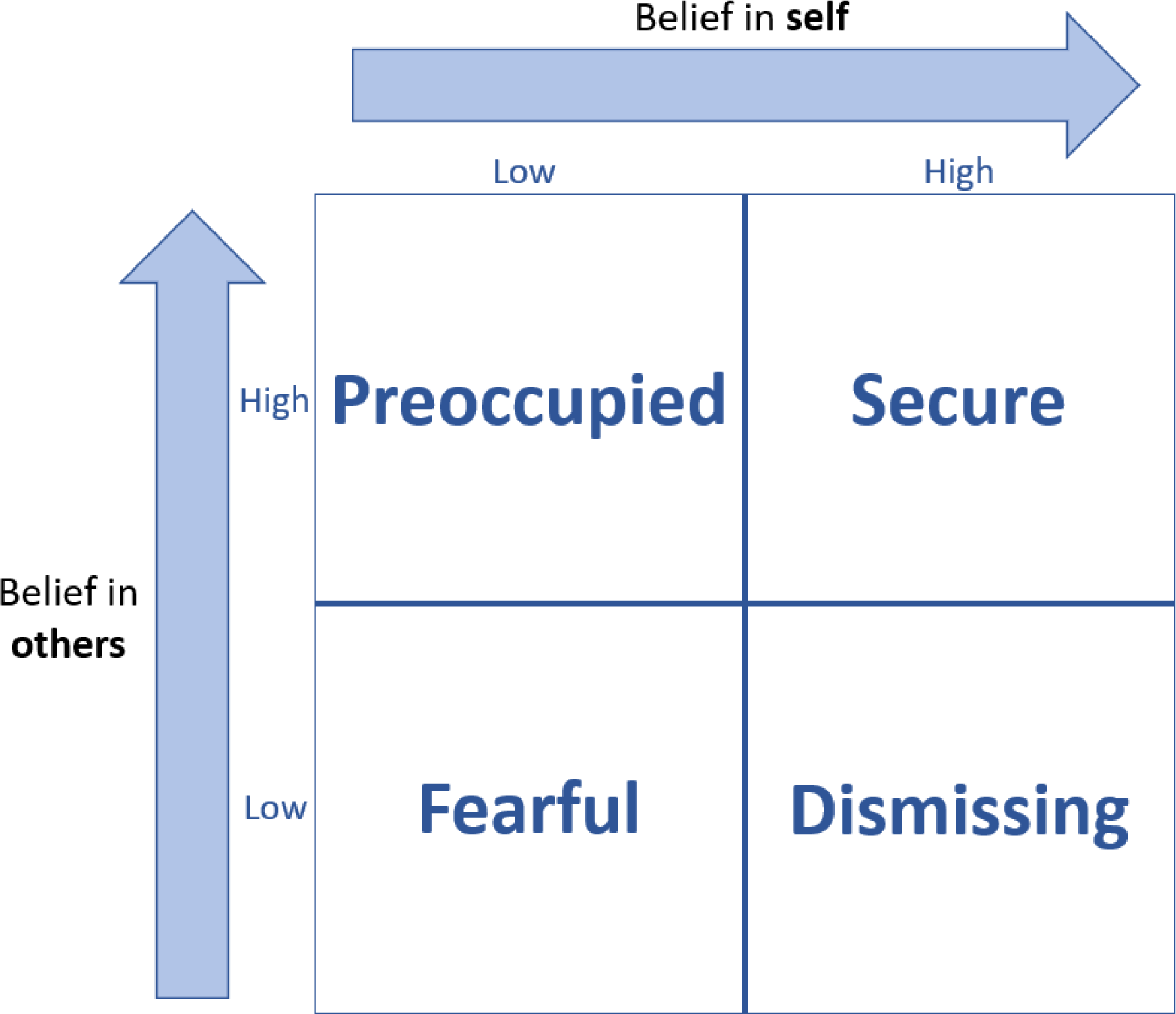
The four adult attachment styles based on the model of adult attachment described in Bartholomew and Horowitz (1991) ^5^. In terms of the attachment dimensions of anxiety and avoidance, a low belief in self represents a high anxiety over abandonment whereas a low belief in others represents high avoidance of attachment.

The attachment system is a psychobiological model for the activation of a matrix of brain areas in response to a perceived threat ^36, 63^. The activation of these brain areas serves to seek safety for self-preservation ^36, 42^. Interestingly, many of the brain regions that govern safety-seeking behaviours in response to a threat, overlap with brain areas that are activated during the pain experience, and during experience of placebo analgesia ^60^. It therefore follows to question whether those who struggle to find a true sense of safety in stressful situations (that is, insecurely attached individuals, with a low belief in themselves to cope, and/or a low belief in others to support them) may be more prone to developing chronic pain. Further, there is observational evidence that cohorts with chronic pain have a larger proportion of individuals with insecure attachment than national samples ^4, 12, 15, 32^. Along the same lines, in a survey of 1361 British people registered at three general practices, those with chronic widespread pain were 70% more likely to have an insecure attachment than pain-free individuals ^15^.

No study to-date has measured attachment style in a general population, followed by an investigation into an association with chronic pain. As such, we explored the social aspect of pain through adult attachment with a view to identifying those at greater risk of developing pain, and who may have a worse experience. Thus, in this study we conducted a large online survey of a general South African population to determine if prevalence of chronic pain associated with adult attachment style, and secondly, in those with chronic pain, whether pain intensity and interference associated with attachment style.

## Methods

### Ethical clearance

The study protocol was approved by the University of the Witwatersrand’s Human Research Ethics Committee (Medical), which adheres to the principles of the Declaration of Helsinki and the Declaration of the World Medical Association (clearance certificate number: M210449).

### Participants

Male and female individuals of 18 years old and above, who were born in, and currently residing in, South Africa, were invited to participate in an online survey. We distributed the survey introduction and link via social media platforms and university emails at the University of the Witwatersrand, University of the Free State, and the University of Cape Town, all of which are in South Africa. The survey was conducted between September 2021 and April 2022.

### Survey

The survey consisted of customized questionnaires that assessed demographic and socio-economic factors, including age, race, sex, education, annual household income. Socioeconomic status was inferred by the reported annual household income. Participants were categorized into low (ZAR 1 – ZAR 19 200), middle (ZAR 19 201 – ZAR 307 200) and high income (> ZAR 307 200) categories ^57^. We also assessed psychosocial factors and pain-related variables using other standardized questionnaires. As detailed below, the psychological factors assessed were adult attachment style and stress, anxiety and depression (using the Experience in Close Relationships - Relationship Structures Questionnaire, and the Depression, Anxiety and Stress Scale 21, respectfully). The pain-related variables that were assessed included pain catastrophizing (using The Pain Catastrophizing Scale Questionnaire), and participants who reported having chronic pain, were also required to complete The Brief Pain Inventory – Short form questionnaire to determine the severity of pain, location of pain, number of pain sites, and pain interference.

#### Adult attachment style

The Experience in Close Relationships - Relationship Structures (ECR-RS) Questionnaire was used to assess adult attachment style. The ECR-RS is a 9-item questionnaire with a 6-item subscale for avoidance and a 3-item subscale for anxiety, and has been previously validated in a non-clinical sample ^22^. The 9-items are repeated four times, each with respect to a different relationship (a relationship with a mother/mother-like figure, father/father-like figure, romantic partner, and best friend). The questionnaire is scored on a 7-point Likert scale anchored at 1 (representing “strongly disagree”) to 7 (representing “strongly agree”). The mean scores for the six avoidance subscales and three anxiety subscales for each relationship were calculated, and the mean of these scores over the four relationships gave the global dimensions for attachment anxiety and avoidance ^22^. The global dimensions were then classified into adult attachment styles based on the 4-category model of attachment ^5^. A score ≤ 4 for both attachment anxiety and avoidance dimensions was classified as a secure adult attachment style, while a score of > 4 for both attachment dimensions was classified as a fearful adult attachment style. A score ≤ 4 for the attachment anxiety dimension but >4 for the attachment avoidance dimension was classified as a dismissing adult attachment style. Lastly, a score ≤ 4 for the attachment avoidance dimension and > 4 for the attachment anxiety dimension was classified as a preoccupied adult attachment style. Figure 1 shows the interactions between the dimensions and how attachment anxiety equates to belief in self, and attachment avoidance to belief in others ^5, 42^.

#### Depression, Anxiety and Stress

The Depression, Anxiety and Stress Scale 21 (DASS-21) was used to measure depression, anxiety and stress ^37^. The DASS-21 has been validated in a South African non-clinical population ^16^ and has been found to be an effective measure of depression in patients with chronic pain due to the absence of questions about somatic symptoms ^59^. The DASS-21 is a 21-item questionnaire with three 7-item subscales assessing depression, anxiety and stress. The questionnaire was scored on a 4-point Likert scale anchored at 0 (representing “never” applies to the individual) to 3 (representing “almost always” applies to the individual). The total score for each subscale was 21, with scores above 13, 9 and 16 indicating extremely severe depression, anxiety and stress, respectively.

#### Pain catastrophizing

The Pain Catastrophizing Scale (PCS) questionnaire was used to measure the participant’s tendency to adopt pain catastrophizing thoughts. The PCS is a 13-item questionnaire assessing pain rumination, pain magnification and helplessness, and has previously been validated in a non-clinical sample ^58^. The PCS was scored on a 5-point Likert scale anchored at 0 (representing “not at all”) to 4 (representing “all the time”). The highest possible score for the PCS questionnaire is 52 ^58^.

#### Chronic pain

Chronic pain was classified as pain experienced most days for the last three months ^61^. Participants who were classified as having chronic pain, answered The Brief Pain Inventory – Short form (BPI-sf) questionnaire ^13^, which was used to determine pain severity, location of pain, number of pain sites, and pain interference. The questions were scored on a numerical rating scale (NRS) anchored at 0 (no pain) to 10 (worst pain imaginable). A total pain severity score was calculated by finding the mean of the reported pain intensities of the participants’ worst, least, average and current pain over the last week ^9^. Similarly, the pain interference score was calculated by finding the mean of reported interference of pain in general activity, mood, walking, work, relations with people, sleep, and enjoyment of life ^9^.

### Data analysis

Before determining whether chronic pain was associated with attachment style, we first wanted to ensure we had sufficient coverage of all attachment styles in the survey. Based on the prevalence of adult attachment styles in European and North American populations, where the least frequent attachment style (Preoccupied attachment style) had a prevalence ranging from 9% to 13 % ^4, 14^, a power analysis determined that a minimum sample size of 2305 was required to answer the question of whether adult attachment style is associated with chronic pain prevalence in a general South African population with a 95% confidence level of certainty.

Descriptive data are reported as total number (N) and percentages (%) for categorical data and median [interquartile range (IQR)] for skewed continuous data. We used variations of generalized linear models for our inferential analyses. For our primary analysis (the relationship between adult attachment style and chronic pain prevalence), a univariate logistic regression model was run for adult attachment style with chronic pain, using incidence rate ratios to describe the relationship. Thereafter, a multivariate logistic regression model was run to control for covariate and confounding factors for chronic pain, excluding collider variables. A subsequent multivariate mediation analysis was also performed to assess the direct effect of adult attachment style on chronic pain. For our secondary analysis looking at the relationship between adult attachment style and the measures of the burden of chronic pain (chronic pain severity, interference and number of pain sites), univariate linear regression models (adult attachment with pain severity, interference or pain sites) were run. Subsequently, multivariate linear regression models (including covariates and confounders) and multivariate mediation analyses were run for each measure of the burden of pain. Lastly, a dropout analysis was performed to compare the attachment styles, gender, annuals household income, age, depression, anxiety and pain catastrophizing of individuals who reported chronic pain but did not complete the BPI and those who reported chronic pain and completed the BPI. The dropout analysis was for the purpose of identifying possible reasons for lack of completion of the BPI questionnaire.

Fig. 2 illustrates the different types of variables to explain why variables were included or excluded in the multivariate models. Data are reported as odds ratios and crude estimates (95% confidence intervals [CI]) for the logistic regressions, incidence rate ratios and crude estimates (95% [CI]) for Poisson regressions and estimates with 95% [CI] for linear regressions. All data processing and analysis was performed in the R statistical environment (v4.2.1) ^49^, using the following packages: DHARMa ^26^, emmeans ^34^, lme4 ^6^, pscl ^28^, psych ^53^, sjPlot ^38^, tidyverse ^66^ and VGAM ^69^. A p-value<0.05 was considered to be statistically significant for all analyses. All data, analysis scripts and analysis script outputs are available at Figshare: https://doi.org/10.6084/m9.figshare.23531076.v1.

**Fig. 2:**
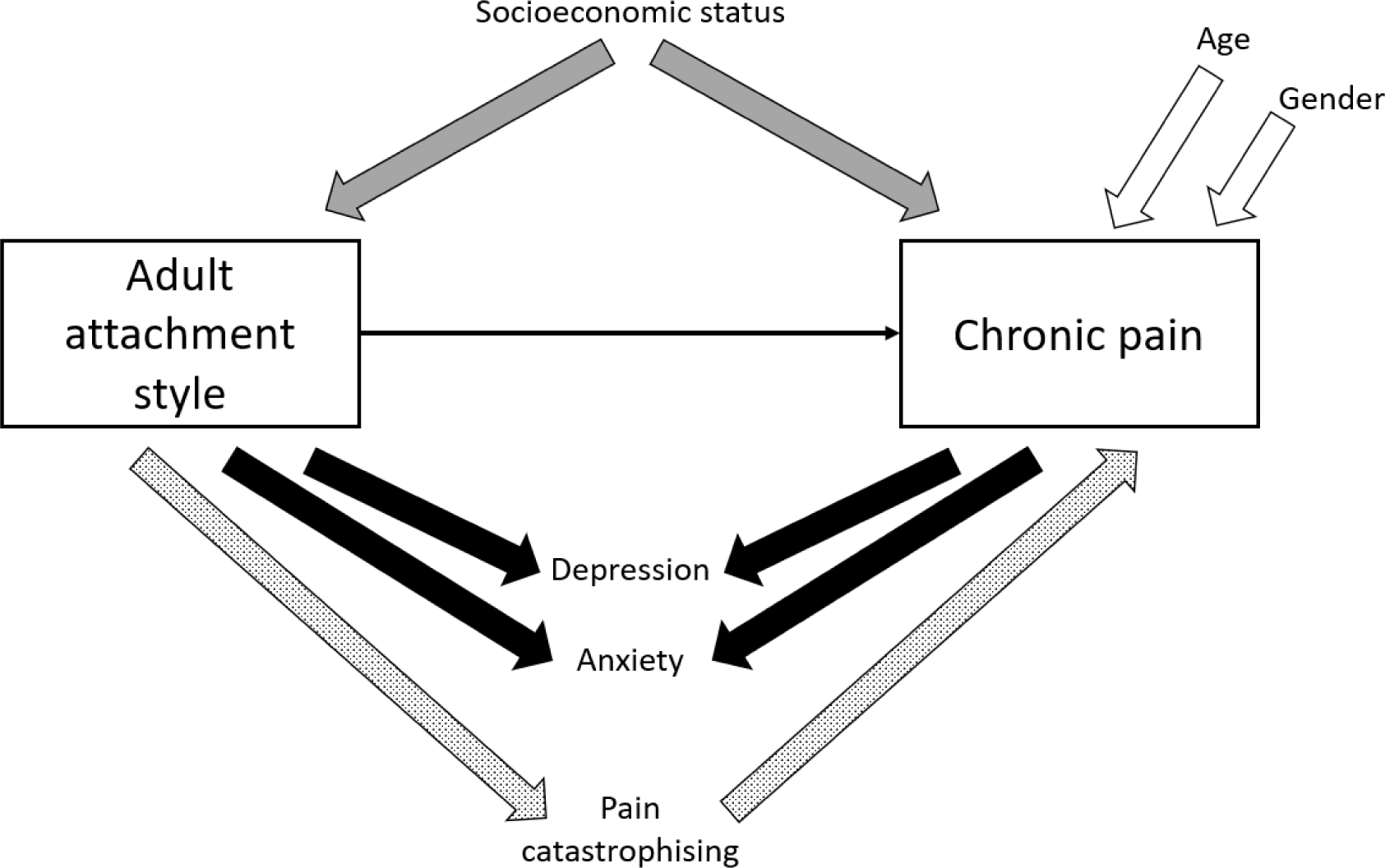
Types of variables encountered in our study, and how each of them were accounted for in our multivariate analyses. Age and gender have been previously found to affect chronic pain (white arrows), making them *covariate factors* ^30^. Socioeconomic status has been found to affect adult attachment style ^54^ and chronic pain ^48^ (grey arrows), making it a *confounding factor*. To determine the total effect of the independent variable (adult attachment style) on the outcome variable (chronic pain), both confounders and covariates should, and were, accounted for in a multivariate analysis. Adult attachment style and chronic pain have both been found to affect depression and anxiety ^27, 51, 68^ (black arrows), making them *collider variables*. A collider should, and was, removed from multivariate models. Lastly, adult attachment style has been found to affect pain catastrophizing ^40^, which in turn, has been found to affect chronic pain ^56^ (pattern arrow), making it a *mediating factor*. Mediators should also be excluded from a model when looking for the total effect of the independent variable on the outcome variable. However, mediators should be included when looking for a direct effect of the independent variable of the outcome variable (a mediation analysis) ^39^.

## Results

We had 3356 survey entries, of which 2371 individuals completed the entire survey and were included in the analyses. The age range of the participants was 18 to 80 years old with a median (IQR) of 23 (20-28). At least secondary education had been completed by 99% (2362/2371) of the participants, and 79% (1877/2371) of the participants had an annual household income of greater than R19 200.00 (middle and high income households ^57^). The prevalence of chronic pain was 27% (635/2371) in our sample, and more details of our cohort are described in Table 1.

**Table 1:** A summary of the demographic data and adult attachment style proportions.

| Characteristic | Final sample N (%) |
| --- | --- |
| Gender |  |
| Men | 596 (25) |
| Women | 1747 (74) |
| Other | 28 (1) |
| South African Province |  |
| Eastern Cape | 53 (2) |
| Free State | 382 (16) |
| Gauteng | 1277 (54) |
| Kwa-Zulu Natal | 113 (5) |
| Limpopo | 43 (2) |
| Mpumalanga | 37 (1) |
| Northern Cape | 14 (1) |
| North West | 28 (1) |
| Western Cape | 424 (18) |
| Race/Ethnicity |  |
| Asian | 18 (1) |
| Black | 956 (40) |
| Indian | 161 (7) |
| Mixed race | 119 (5) |
| White | 1062 (45) |
| Other | 55 (2) |
| Annual household income |  |
| No income | 115 (5) |
| Low income | 379 (16) |
| Middle income | 1030 (43) |
| High income | 847 (36) |
| Adult attachment style* |  |
| Secure | 1 824 (76.93) |
| Dismissing | 246 (10.38) |
| Preoccupied | 172 (7.25) |
| Fearful | 129 (5.44) |

\*Adult attachment style was assessed using the Experience in Close Relationships - Relationship Structures (ECR-RS) Questionnaire<sup>22</sup>.

Table 2 summarizes the univariate associations between variables measured (including adult attachment style) and prevalence of chronic pain. We found that increased prevalence of chronic pain is associated with women, insecure adult attachment styles, lower household income, and increased depression, anxiety and pain catastrophizing. The reference variables for the univariate logistic regression models were “Men” (for gender), “Secure” (for the adult attachment style) and “No income” (for annual household income).

**Table 2:**
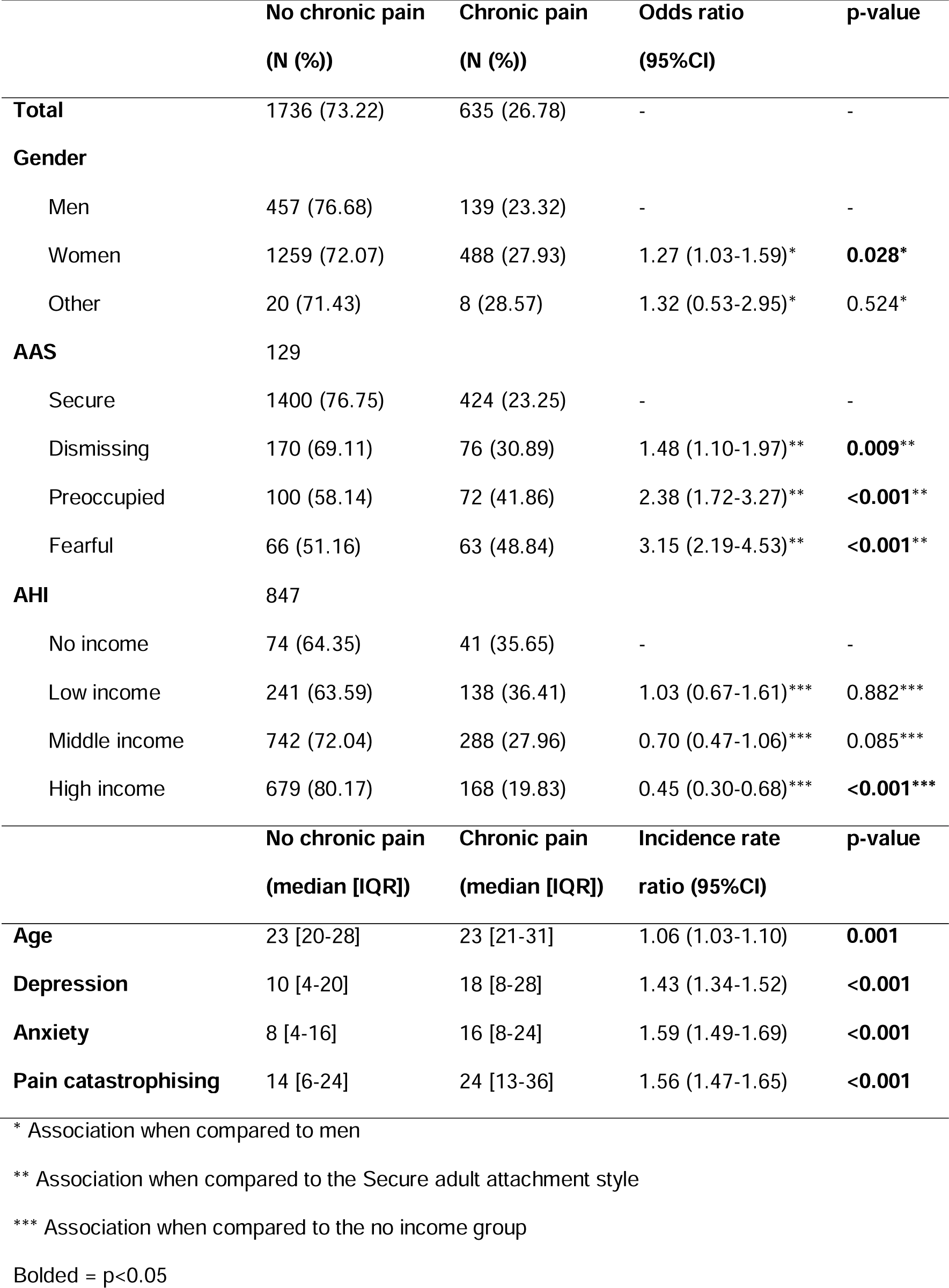
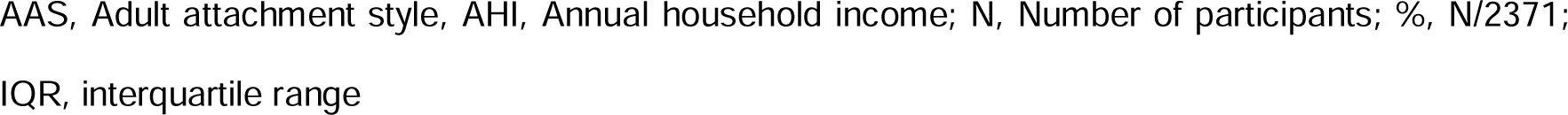
Factors associating with the presence of chronic pain in the 2371 survey.

|  | **No chronic pain**<br>**(N (%))** | **Chronic pain**<br>**(N (%))** | **Odds ratio**<br>**(95%CI)** | **p-value** |
| --- | --- | --- | --- | --- |
| **Total** | 1736 (73.22) | 635 (26.78) | - | - |
| **Gender** |  |  |  |  |
| Men | 457 (76.68) | 139 (23.32) | - | - |
| Women | 1259 (72.07) | 488 (27.93) | 1.27 (1.03-1.59)* | **0.028\*** |
| Other | 20 (71.43) | 8 (28.57) | 1.32 (0.53-2.95)* | 0.524* |
| **AAS** | 129 |  |  |  |
| Secure | 1400 (76.75) | 424 (23.25) | - | - |
| Dismissing | 170 (69.11) | 76 (30.89) | 1.48 (1.10-1.97)** | **0.009\*\*** |
| Preoccupied | 100 (58.14) | 72 (41.86) | 2.38 (1.72-3.27)** | **<0.001\*\*** |
| Fearful | 66 (51.16) | 63 (48.84) | 3.15 (2.19-4.53)** | **<0.001\*\*** |
| **AHI** | 847 |  |  |  |
| No income | 74 (64.35) | 41 (35.65) | - | - |
| Low income | 241 (63.59) | 138 (36.41) | 1.03 (0.67-1.61)*** | 0.882*** |
| Middle income | 742 (72.04) | 288 (27.96) | 0.70 (0.47-1.06)*** | 0.085*** |
| High income | 679 (80.17) | 168 (19.83) | 0.45 (0.30-0.68)*** | **<0.001\*\*\*** |
|  | **No chronic pain**<br>**(median [IQR])** | **Chronic pain**<br>**(median [IQR])** | **Incidence rate**<br>**ratio (95%CI)** | **p-value** |
| **Age** | 23 [20-28] | 23 [21-31] | 1.06 (1.03-1.10) | **0.001** |
| **Depression** | 10 [4-20] | 18 [8-28] | 1.43 (1.34-1.52) | **<0.001** |
| **Anxiety** | 8 [4-16] | 16 [8-24] | 1.59 (1.49-1.69) | **<0.001** |
| **Pain catastrophising** | 14 [6-24] | 24 [13-36] | 1.56 (1.47-1.65) | **<0.001** |

\* Association when compared to men
\*\* Association when compared to the Secure adult attachment style
\*\*\* Association when compared to the no income group
Bolded = $p < 0.05$
AAS, Adult attachment style, AHI, Annual household income; N, Number of participants; %, N/2371;
IQR, interquartile range

Table 3 shows the results of two multiple logistic regressions of the association between adult attachment style and chronic pain, with Secure as the reference attachment style. The first model investigates a total effect of adult attachment on chronic pain. The second model (“Direct effect”) is a mediation analysis showing the mediating effect of pain catastrophizing on the relationship between adult attachment style and chronic pain, including confounders (annual household income), covariate (age and gender), but excluding the colliders (depression and anxiety) and mediators (pain catastrophizing).

**Table 3:**
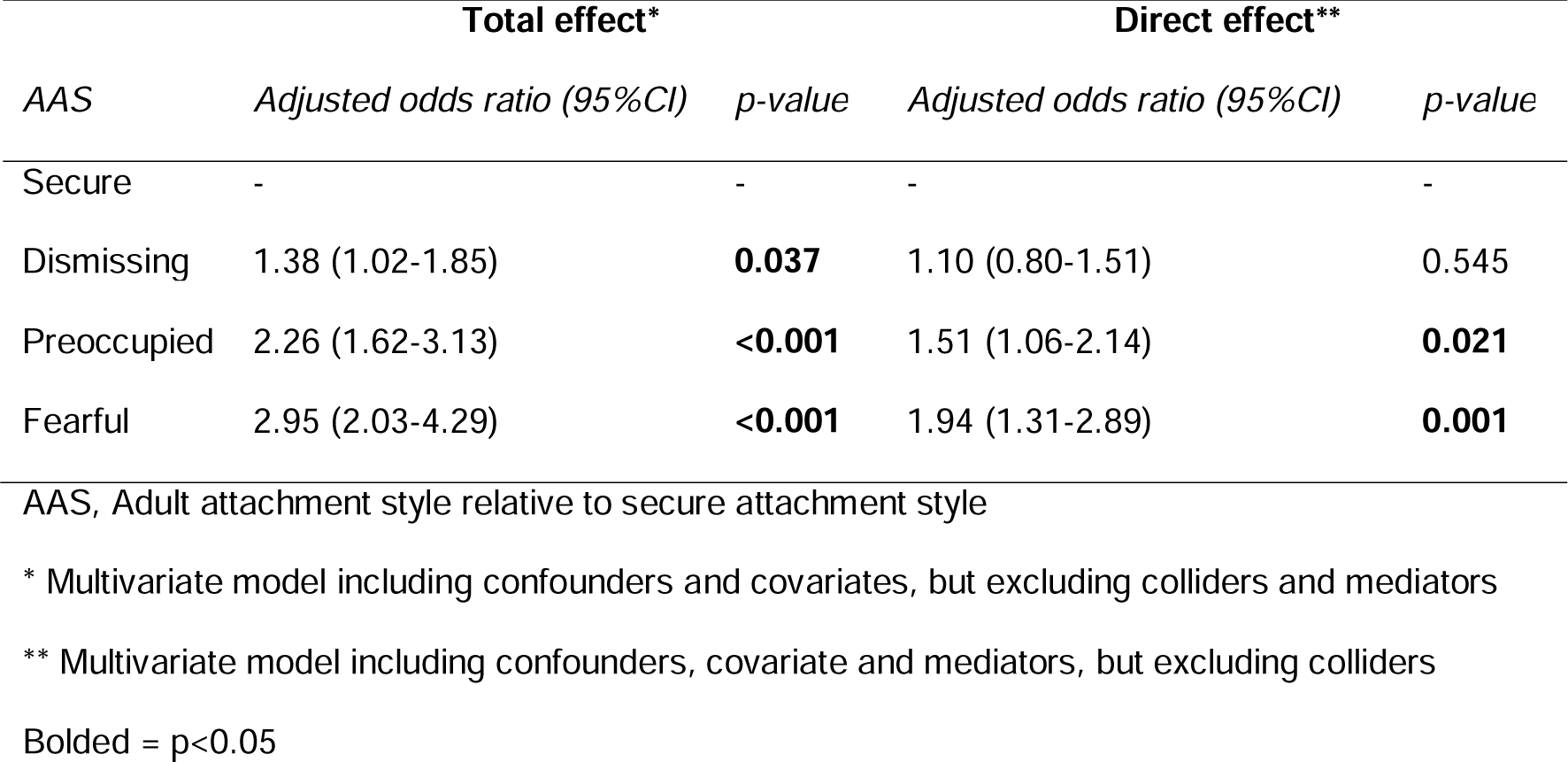
Multivariate analyses showing the total and direct effect of adult attachment style on chronic pain is our total sample (N = 2371).

| AAS | Total effect* |  | Direct effect** |  |
| --- | --- | --- | --- | --- |
|  | *Adjusted odds ratio (95%CI)* | *p-value* | *Adjusted odds ratio (95%CI)* | *p-value* |
| Secure | - | - | - | - |
| Dismissing | 1.38 (1.02-1.85) | **0.037** | 1.10 (0.80-1.51) | 0.545 |
| Preoccupied | 2.26 (1.62-3.13) | **<0.001** | 1.51 (1.06-2.14) | **0.021** |
| Fearful | 2.95 (2.03-4.29) | **<0.001** | 1.94 (1.31-2.89) | **0.001** |

AAS, Adult attachment style relative to secure attachment style
\* Multivariate model including confounders and covariates, but excluding colliders and mediators
\*\* Multivariate model including confounders, covariate and mediators, but excluding colliders
Bolded = $p < 0.05$

### Secondary analysis

The secondary analysis explored the burden of pain (through number of pain sites, pain severity and pain interference) for different attachment styles using the results of the Brief Pain Inventory. Two hundred and eight individuals were classified as having chronic pain but did not complete the BPI. Thus, only 427 individuals (those with chronic pain who completed the BPI questionnaire) were included in the secondary analysis. A dropout analysis showed the following associations with those that had dropped out: these individuals were more likely to be male (compared to female) (Odds ratio [CI], 0.64 [0.43 – 0.94], p=0.024) with significantly lower depression (Odds ratio [CI], 0.97 [0.96 – 0.99], p<0.001), anxiety (Odds ratio [CI], 0.96 [0.94 – 0.97], p<0.001) and pain catastrophizing (Odds ratio [CI], 0.99 [0.98 – 1.00], p=0.037).

The pain severity and pain interference scores for the adult attachment styles are summarized in Fig. 3. Two separate univariate linear regression models were run for adult attachment with pain severity score and pain interference score. Compared to the secure attachment style, the dismissing (Estimate [95%CI] = 0.48 [0.00-0.96], p-value = 0.049) and fearful (Estimate [95%CI] = 0.88 [0.36-1.40], p-value = 0.001) attachment styles significantly predicted increased pain severity scores. Pain interference scores were significantly higher in Dismissing (Estimate [95%CI] = 0.70 [0.04-1.37], p-value = 0.038), Preoccupied (Estimate [95%CI] = 0.93 [0.23-1.63], p-value = 0.010) and Fearful (Estimate [95%CI] = 1.31 [0.59-2.03], p-value<0.001) attachment styles when compared to Secure attachment. Multivariate analyses were run to determine the total and direct effect of adult attachment style on pain severity and pain interference score. The Fearful adult attachment style was associated with a total effect of increased pain severity score when compared to the Secure attachment (Estimate [95%CI] = 0.78 [0.26-1.30], p = 0.004). After a mediation analysis to account for pain catastrophizing, we found that pain catastrophizing mediated the relationship between attachment style and pain severity, resulting in no direct associations between adult attachment style and pain severity score. There was a total effect of all three insecure attachment styles being associated with worse pain interference scores when compared to the Secure attachment style (Dismissing: Estimate [95%CI] = 0.73 [0.06-1.41], p = 0.032; Preoccupied: Estimate [95%CI] = 0.88 [0.17-1.58], p = 0.015; Fearful: Estimate [95%CI] = 1.17 [0.44-1.90], p = 0.002). After running a mediation analysis to account for pain catastrophizing, pain catastrophizing mediated the relationship between adult attachment and pain interference, resulting in no direct associations.

**Fig. 3:**
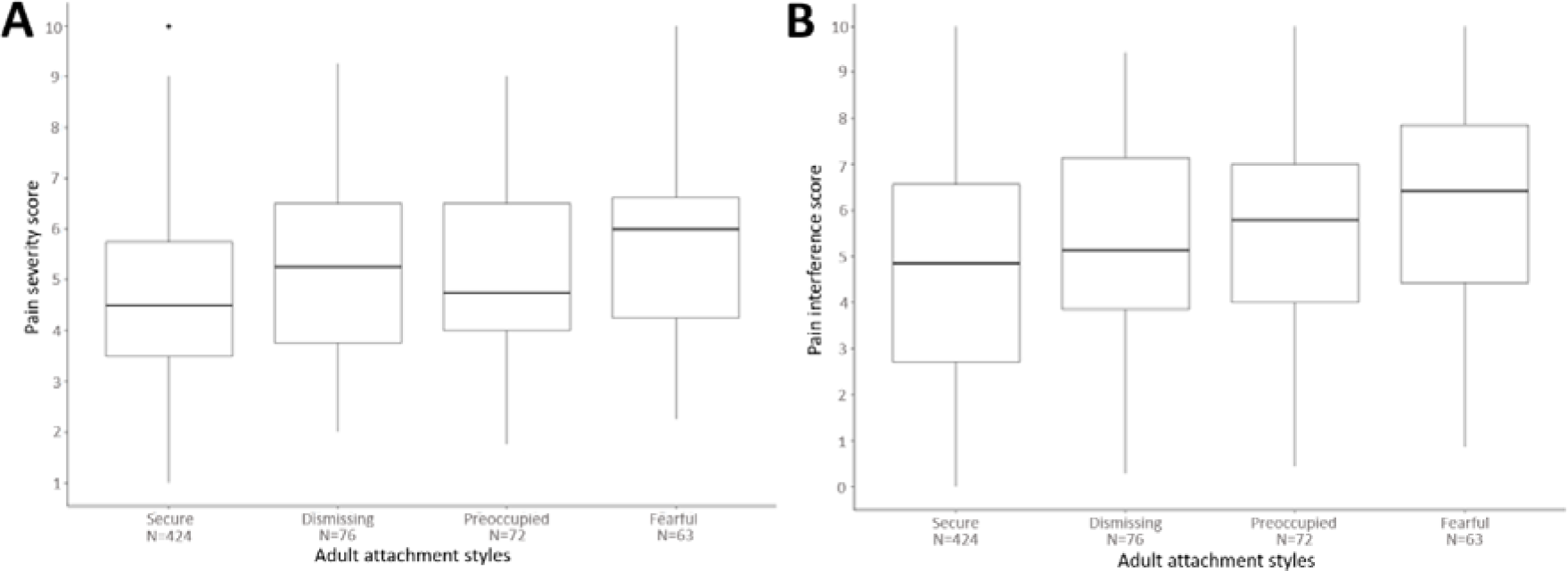
Pain severity score (Panel A) and Pain interference score (Panel B) in the last week for different adult attachment styles. Only individuals with chronic pain who completed the BPI were included (N=427).

With regards to number of painful body sites in individuals reporting chronic pain, a univariate analysis showed that individuals with a Preoccupied attachment had 1.32 times more pain sites (95% CI: 1.10-1.56, p=0.002) and individuals with Fearful attachment had 1.30 times more pain sites (95% CI: 1.09-1.55, p=0.003) than individuals with Secure attachment. Individuals with dismissing attachment did not have a difference in the number of pain sites compared to the securely attached individuals. After running multivariate analyses for the number of pain sites across the various attachment styles, there was a total effect whereby the Fearful adult attachment style was associated with 0.78 times more pain sites compared to the Secure attachment style (95%CI: 0.26-1.30, p = 0.003). After running a mediation analysis that accounted for pain catastrophizing, pain catastrophizing also mediated the relationship and no direct effects of the adult attachment styles on number of pain sites were found.

## Discussion

We conducted a survey of over 2000 individuals to determine associations between adult attachment style and prevalence of chronic pain. In our young, well-educated and primarily female (74%) cohort with predominantly a middle-to-high socioeconomic status, depression, anxiety and pain catastrophizing were increased in those with, compared to those without, chronic pain. Moreover, we found that insecure attachment styles were associated with an increased prevalence of chronic pain. Both preoccupied and fearful styles were directly associated with prevalence of chronic pain, and of note, the prevalence of chronic pain was more than double in individuals with a fearful attachment style compared to securely attached individuals. The relationship between adult attachment style and measures of burden of pain (pain intensity, interference and number of pain sites) was mediated by pain catastrophizing in those with chronic pain. In other words, adult attachment style was not directly associated with the burden of chronic pain, although the effect of pain catastrophizing as a mediator was evident. Our data contribute to the emerging idea that a relationship exists between attachment style and chronic pain. We provide the first report that attachment style in a general population is associated with chronic pain prevalence, while pain catastrophizing mediates the relationship between attachment and the burden of chronic pain.

Chronic pain prevalence is typically low in young people, for example, previously published data in a national sample of South Africans have demonstrated that 13% of 24-34 year olds have chronic pain ^30^. In that same study ^30^, the prevalence of chronic pain in women was 20% across all age groups, which is of note because pain is more prevalent in women than in men ^50^, and 74% of our cohort was female. In our study, the prevalence of chronic pain was 23% in the securely attached individuals, but 39% in individuals with insecure attachment styles. Indeed, specifically within the insecurely attached individuals, those with a fearful attachment had a 49% prevalence of chronic pain. What is remarkable is that this association with chronic pain, and these high prevalence rates of chronic pain were evident in a rather young cohort, with an average age in the early twenties (median age 23 years; IQR 20-28).

The association with insecure attachment and chronic pain prevalence makes sense given the overlapping neurocircuitry between physical and social pain ^18, 19^ and between experimental pain and expectations of social safety or threat ^20^. Essentially, there is evidence that when one perceives an environment and/or stimulus as threatening, the intensity of pain is increased ^31, 62, 64^. Furthermore, there are associations between insecure attachment styles and factors associated with pain, including poorer mental health, greater number of sleep disruptions and greater levels of inflammation ^1, 24, 29^, all of which impact the perception of pain ^3^. Whilst these findings suggest that an association between having chronic pain and insecure attachment style makes logical sense, studies exploring this association and explicitly looking at potential mechanisms are lacking.

One of the issues with attachment and pain literature to date is the heterogeneity of study designs, including the reporting of attachment using secure vs. insecure, the two attachment dimensions (attachment anxiety and attachment avoidance), or the four-category model, as we have done here. Whilst using the two dimensions may have more statistical power ^23^, it loses the nuance of the overlap of both scales; for example, having both high attachment anxiety (low belief in self) and high attachment avoidance (low belief in others). Based on our results, we hypothesize that there may be an additive effect of having high attachment anxiety *and* avoidance, that is, a fearful attachment style, compared to just one high dimension, as is the case for preoccupied and dismissing styles. This hypothesis requires further investigation.

Recent work suggests that attachment style is modifiable ^21, 25^ and so even if an individual develops an insecure attachment during childhood, a secure attachment may be earned through close, supportive relationships or psychological therapy ^43, 55^. On the other hand, there is some evidence from a 20-year longitudinal study of 50 individuals, that stressful life events later in life may lead to the *loss* of a secure attachment style obtains in infancy ^65^. However, at this stage many questions remain unclear, including whether chronic pain may also act as one of those stressful life events that leads to loss of secure attachment (and all its associated benefits, including improved emotional regulation and decreased physiological stress ^35, 44, 47^). The somewhat opposing cause-effect question also remains unclear. In other words, if one develops an insecure attachment style from childhood interpersonal relationships, does that predispose one to the development of chronic pain?

Longitudinal studies are required to answer such questions. Such questions have important implications on treatment options, response to treatment, progression of disease, and overall quality of life in patients with chronic pain. Indeed, preliminary evidence suggests that individuals with insecure attachment have lower adherence rates to pain management programmes and poorer psychological outcomes following pain management programmes ^2, 10, 11, 32^. These data suggest that profiling the attachment of patients, and a better understanding of the different responses to management programmes according to attachment security, may path the way forward to personalized treatment options with more favourable clinical outcomes.

We found a statistically significant total effect difference in pain intensity, and particularly pain interference, between attachment styles, which was mediated by pain catastrophizing. However, the increase in the severity and interference scores for the insecure attachment styles were not clinically significant (less than 1.2 units (11%) difference between the scores of the secure attachment style and the insecure attachment styles) ^17^. The lack of a clinically-relevant association between attachment style and pain intensity and disability in those with chronic pain has been reported previously ^2, 15, 32, 41^. In a cohort similarly-sized to ours, with chronic widespread pain, insecure attachment styles associated with greater number of pain sites when compared to the secure attachment style ^15^. We found a similar association where the number of pain sites was associated with adult attachment style, although this relationship was also mediated by pain catastrophizing. The role of pain catastrophizing as a mediator between adult attachment and measures of the burden of pain was highlighted in our study. The significant changes in the burden of chronic pain measures for insecure attachment style prior to the mediation analysis were not clinically relevant. However, when these statistically significant increases in these chronic pain burden measures fell away following the mediation analyses, the importance of studying pain catastrophizing in the context of adult attachment and chronic pain becomes evident.

There were limitations to our study. Our study was conducted in 2021 during the COVID-19 pandemic, and general depression and anxiety increased during this period ^7, 67^. As such, depression and anxiety rates in our cohort may have been higher than if we had measured the year before, but attachment styles likely would have remained the same. We recruited a convenience sample, that is, a sample of individuals responding to our social media or university email advert to complete a survey on attachment style and pain. The respondents were 75% female, 99% had completed at least secondary education and all had access to a device and data with which to respond. This sex, education and socioeconomic profile is not representative of the South African population ^57^ but rather makes our data more generalizable to developed countries where there are increased numbers of individuals who are educated with a higher socioeconomic status ^45^. Another limitation was that 33% (208/635) of the individuals with chronic pain did not complete the Brief Pain Inventory and so their pain intensity and interference data were missing. It is not clear why this may have been the case. The dropout analysis indicated that in the 208 individuals with chronic pain who did not complete the Brief Pain Inventory questionnaire, these individuals were more likely to be male.

In this analysis of attachment style and chronic pain in a young and primarily female cohort with a middle-to-high socioeconomic status, not only did we find an association between insecure adult attachment styles and greater prevalence of chronic pain, but chronic pain prevalence was double in individuals with a fearful attachment style when compared to those with a secure attachment style. There were no clinically relevant differences in pain intensity and interference between the attachment styles, although the role of pain catastrophizing as a mediator between adult attachment and the burden of chronic pain was highlighted and warrants further investigation. Our results highlight the need locally to repeat the study in a more nationally-representative sample, and globally, for longitudinal studies to map attachment security as well as incidence and outcomes of chronic pain.

## Data Availability

All data produced are available online at Figshare: 10.6084/m9.figshare.23531076

https://doi.org/10.6084/m9.figshare.23531076.v1

## Acknowledgements

We would like to thank Professor Peter Kamerman for his guidance and advice for the statistical analyses of our data. The study was funded by the University of the Witwatersrand, Faculty of Health Sciences Research Committee. The authors declare no conflicts of interest. All data, analysis scripts and analysis script outputs are available at Figshare: 10.6084/m9.figshare.23531076. The research conducted was not preregistered with an analysis plan in any independent, institutional registry.

